# A phase 2 trial of burosumab for treatment of fibroblast growth factor-23 mediated hypophosphatemia in children and adults with fibrous dysplasia

**DOI:** 10.1101/2025.09.26.25334161

**Authors:** Olivia de Jong, Zubeyir Hasan Gun, Afua Asante-Otoo, Ibrahim Elbashir, Xiaobai Li, Babak Saboury, Vardit Kram, Luis F de Castro, Vivian MacDonald, Alison M Boyce

**Author notes:** Correspondence to: Alison Boyce, MD, Building 30 Room 228 MSC 4320, Bethesda, MD, 20892.

## Abstract

Fibrous dysplasia (FD) is a rare disorder associated with fractures and deformities. FD lesions produce excess phosphaturic hormone fibroblast growth factor 23 (FGF23), leading to hyperphosphaturia in most patients, and hypophosphatemia in those with high FD burden. Skeletal complications are associated with both low-normophosphatemia and frank hypophosphatemia. Burosumab is approved for other forms of FGF23 excess, but there is little evidence to inform use in FD. A phase 2 study investigated the safety and efficacy of burosumab in patients with FD. The primary endpoint was the proportion of participants achieving phosphate levels within a high-normal target range (age and sex-adjusted Z-score –1 to +2). 12 participants (7 children, 5 adults) received burosumab for 48 weeks. Median phosphate Z-score increased from -2.88 (1.65) to 0.22 (1.37), meeting the target in 100% of participants. Alkaline phosphatase levels were elevated at baseline in 8 participants and declined by 49%. PROMIS questionnaires showed trends toward improvements in all domains in children; adult scores showed no identifiable trends. Two children experienced transformational mobility gains, including advancement from full-time wheelchair use to independent ambulation. Lesion biopsies showed no changes in cellularity or composition, and ^18^F-NaF PET/CT scans showed no changes in tracer uptake, suggesting burosumab did not adversely impact lesional activity. Adverse events were mild, and none resulted in treatment withdrawal. Burosumab targeting high-normophosphatemia in patients with FD was well-tolerated, restored phosphate homeostasis, and improved bone turnover. Burosumab has the potential to lead to functional improvements and ambulation gains in severely affected patients and is a valuable tool to reduce the impact of FD-related disability.

## INTRODUCTION

Fibrous dysplasia (FD) is a rare mosaic disorder associated with fractures, pain, and skeletal deformities. It may affect one or multiple bones and may occur in association with skin hyperpigmentation and/or endocrinopathies, termed McCune-Albright syndrome (MAS) [1]. FD/MAS arises due to somatic gain-of-function variants in Gα_s_, resulting in constitutive activation of the G_s_ G-coupled protein receptor. This disrupts differentiation of osteoprogenitor cells, replacing normal bone and marrow with expansile fibro-osseous tissue [2]. The resulting skeletal lesions are comprised of poorly mineralized woven bone, prone to fractures and bowing deformities (Fig 1).

**Figure 1.**
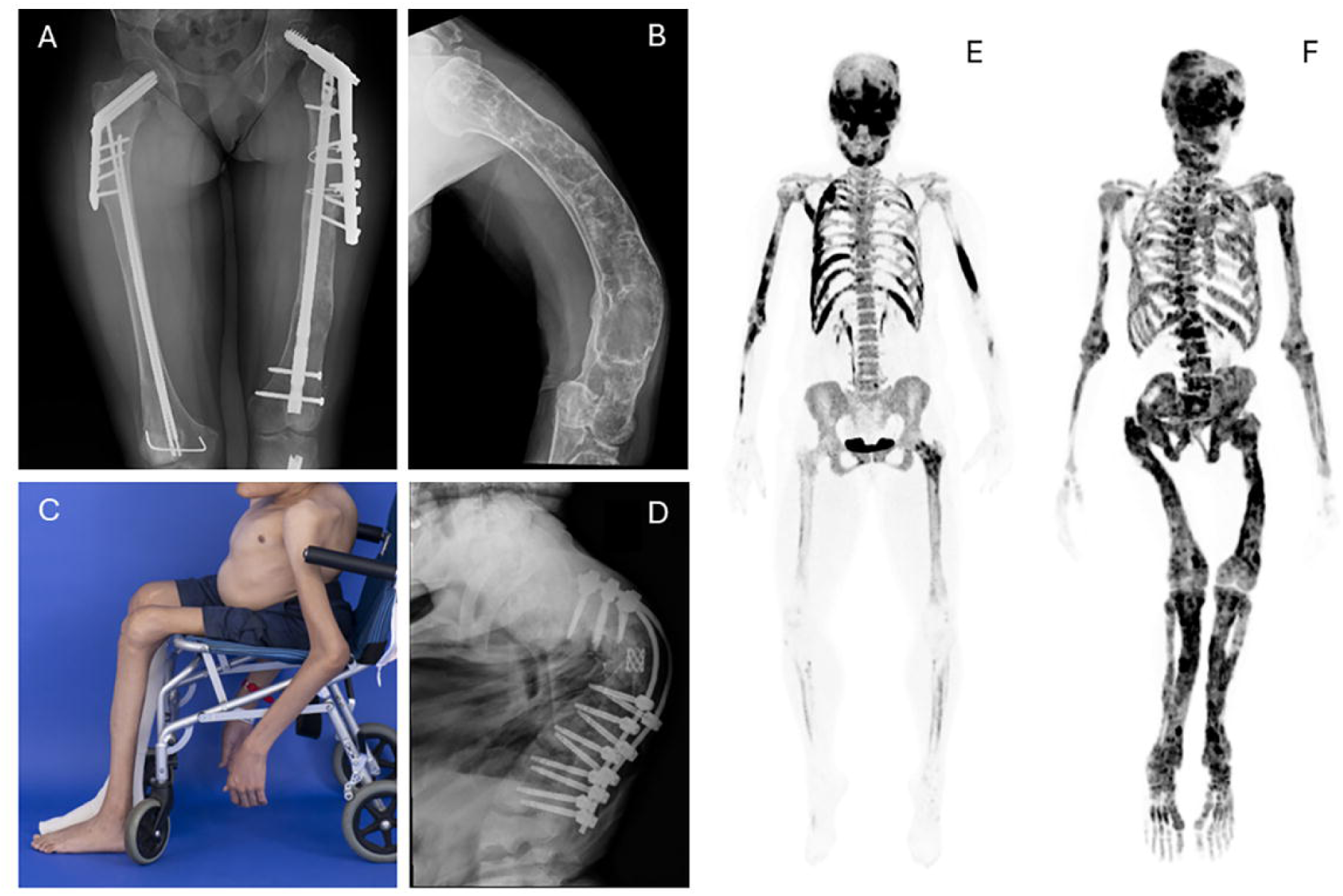
Representative baseline clinical images demonstrating typical features of fibrous dysplasia. A) Bilateral femoral involvement in participant BUR03, with multiple surgical implants for correction of fractures and bowing deformities. B) Humerus radiograph in participant BUR01 reveals severe bowing deformity. C) Photograph of participant BUR07 demonstrates severe scoliosis, and sequelae of long-term mobility impairment including muscle wasting and joint contractures in the upper and lower extremities. D) Spinal radiograph for this participant demonstrates severe curvature with fixation device present. The right-hand panels include representative ^18^F-NaF PET/CT scans demonstrating the spectrum of disease severity for participants in this trial. Fibrous dysplasia involvement is indicated by dark areas of tracer uptake. E) The most mildly affected participant (BUR11) presents with fibrous dysplasia throughout the skull, ribs, upper extremities, and left femur, with a Skeletal Burden Score of 28. F) The most severely affected participant (BUR02) has panostotic fibrous dysplasia with a Skeletal Burden Score of 75.

Excessive FGF23 production from Gα_s_-variant bearing osteoprogenitor cells results in renal phosphate wasting in most patients, and frank hypophosphatemia in a subset of patients with high skeletal FD burden [3]. Because FD tissue is inherently dysplastic and poorly mineralized, it is at high risk for deleterious effects of hypophosphatemia. Retrospective studies have demonstrated that a diagnosis of hypophosphatemia is associated with an increased risk of FD-related fractures and deformities [4-7]. Previous work from our group has demonstrated a linear relationship between serum phosphate levels and FD-related complications, identifying that fractures and surgeries are lowest in patients who have phosphate levels at the mid-to-upper portion of the normal range [8]. This suggests that even mild degrees of hypophosphatemia may increase the risk of FD-related complications, and that patients may benefit from maintaining phosphate levels in the mid to upper part of the normal range.

Like other disorders of FGF23 excess, management of hypophosphatemia in FD has traditionally focused on repletion with oral phosphate and active vitamin D analogs. However, this regimen is complicated by dose-limiting side effects, including gastrointestinal intolerance and renal toxicity [9]. Normalization of serum phosphate is not a practical goal with conventional treatment, placing patients at continued risk of skeletal complications from persistent hypophosphatemia. Burosumab, a human recombinant monoclonal antibody to FGF23, represents a newer, targeted treatment approach. Prospective trials have demonstrated safe, efficacious use of burosumab in the FGF23 excess disorders X-linked hypophosphatemia (XLH) and tumor-induced osteomalacia (TIO), leading to improved serum phosphate levels and skeletal outcomes without adverse gastrointestinal or renal side effects [10-15].

Given the known limitations of conventional therapy, burosumab is an intuitive choice for management of hypophosphatemia in FD. However, additional research into its safety and efficacy is needed before it can be used routinely in this population. The potential for FD tissue effects is an important consideration; it is unknown whether burosumab may impact osteoprogenitor cell proliferation or other drivers of lesion growth and activity. Typical clinical management in XLH and TIO involves dosing burosumab to achieve a low-normal phosphate level [16-18], reflecting the target phosphate levels in seminal trials [10-14]. Investigation is therefore needed to determine if targeting a high-normal phosphate level is a safe and effective therapeutic goal in patients with FD.

## METHODS

### Trial Design and Oversight

A phase 2 open-label study was conducted at the NIH (NCT05509595). This investigator sponsored study was supported by Kyowa Kirin, Inc and Ultragenyx, Inc; study design, conduct, and analyses were performed by the investigators. The trial was approved by the NIH Investigational Review Board, and informed consent/assent was obtained from all participants. The study was monitored by a data safety and monitoring committee organized by the National Institute of Dental and Craniofacial Research.

### Participants

Children and adults aged ≥1 year with a confirmed diagnosis of FD were eligible for this trial. Key inclusion criteria included serum phosphate below the 10^th^ percentile for age and sex, in the setting of intact serum FGF23 ≥30 pg/mL [19]. Additional inclusion and exclusion criteria are listed in the protocol (Suppl Appendix).

### Procedures

Participants discontinued conventional treatment for a 2-week washout period. The starting dose of burosumab for pediatric participants was 0.8 mg/kg rounded to the nearest 10 mg for a maximum of 90 mg, administered every 2 weeks. For adults age ≥18 years the starting dose was 0.5 mg/kg rounded to the nearest 10 mg for a maximum of 90 mg, administered every 4 weeks, with the option to increase to every 2 weeks at the discretion of the investigators. Dosing was titrated in 0.2-0.5 mg/kg intervals to achieve a target fasting phosphate level between -1 and +2 standard deviations for age and sex. Age- and sex-adjusted Z-scores were calculated using reference ranges from the CALIPER study, which included thousands of healthy individuals from a multiethnic population [20].

The initial dose of burosumab was administered via subcutaneous injection by a healthcare professional at the NIH Clinical Center. Participants and caregivers were given injection teaching, and subsequent injections were self- or caregiver-administered with telehealth oversight from the study team.

Fasting laboratory tests were obtained prior to administering burosumab. Phosphate, 1,25-dihydroxyvitamin D, 25-hydroxyvitamin D, parathyroid hormone, tubular maximum for phosphate reabsorption per glomerular filtration rate (TmP/GFR), alkaline phosphatase, osteocalcin, procollagen-1-propeptide (P1NP), and C-telopeptide (CTX) levels were assessed throughout the trial, with details available in the protocol (Suppl Appendix). Labs were drawn at the NIH Clinical Center at baseline, 24, and 48 weeks, and used for efficacy and safety analyses. Between NIH visits, labs were drawn at participants’ local laboratories; these levels were used for safety analyses and dose adjustments.

Functional mobility was assessed by manual muscle testing, range-of-motion assessments, and 9-Minute Walk test. Patient-reported outcomes included PROMIS questionnaires related to Pain Intensity (Pediatric and Parent Proxy version 1.0, Adult version 2.0), Pain Interference (Pediatric and Parent Proxy v 2.0, Adult v 1.1), Mobility (Pediatric and Parent Proxy version 2.0, Adult Mobility Lower Extremity v 1.0), and Fatigue (Pediatric and Parent Proxy v 2.0, Adult FACIT 13a v1.0). Pediatric questionnaires were administered to children age 8-17 years, and parent proxy questionnaires were administered to caregivers of children ≤7 years. Adult participants were administered the 36-item Short Form Health Survey, and pediatric participants were administered the SF-10 Child Health Survey.

Quantitative assessment of functional and patient-reported outcomes in FD/MAS is impacted by multiple challenges, including disease rarity, high levels of disability, and broad clinical heterogeneity between patients. To address these challenges, a qualitative Activities of Daily Living Questionnaire was developed to evaluate individualized changes after initiation of burosumab (Suppl Appendix). The questionnaire was developed by investigators at NIH, Ultragenyx, and Kyowa Kirin, and informed by a Patient and Public Involvement exercise conducted with adult patients and caregivers in the United States and Europe [21]. Participants and/or caregivers were asked at the baseline visit to provide 3 activities of daily living that were affected by FD at home or school/work (e.g., mobility, climbing stairs, dressing, playing with peers, catching a bus, etc) and to estimate the extent of impact upon their activity level. These activities were then reassessed at weeks 24 and 48 to determine the change in FD-related impact.

Key safety assessments included evaluation of lesion activity. ^18^F-NaF PET/CT scans were performed at baseline and 48 weeks to quantify radiographic changes [22 23]. FD lesion biopsies were performed in adult participants at baseline and 48 weeks to evaluate changes in lesional cellularity and composition. Biopsy sites were chosen jointly by the investigators and surgeons to ensure that the procedure was minimally invasive, and biopsies were performed in the interventional radiology suite at the NIH Clinical Center using core needles under CT guidance. The same sites were biopsied before and after burosumab treatment.

All adverse events were recorded and graded according to the Common Terminology Criteria for Adverse Events (CTCAE) Version 5.0. Metabolic panels, including calcium, parathyroid hormone, and 24-hour urinary calcium excretion, were performed at regular intervals. Physical examinations, including assessment of body weight and vital signs, were performed at regular intervals. Renal ultrasonography was performed regularly to evaluate for nephrocalcinosis. Anti-burosumab antibodies were evaluated at baseline and end of treatment (Toray Industries, Tokyo, Japan).

### Outcomes

The primary endpoint was the proportion of participants achieving serum phosphate levels within the target range (Z-score -1 to +2) at week 48. Key secondary endpoints included change in bone turnover markers (alkaline phosphatase, osteocalcin, P1NP, CTX) and pharmacodynamic measures (TmP/GFR, 1,25-dihydroxyvitamin D) at 24 and 48 weeks. Patient-reported outcomes and functional assessments were performed using the modalities described above at 24 and 48 weeks. Safety endpoints included treatment-emergent adverse events, which were collected continuously throughout the study through 4 weeks post-burosumab discontinuation. Additional key safety endpoints included change in lesion activity as assessed by ^18^F-NaF PET/CT at baseline and 48 weeks, and change in FD lesion histology and cell proliferation from baseline to 48 weeks.

### Statistical Analysis

Data analyses were conducted in SAS (the SAS Institute, Cary, NC). Changes from baseline at 24 and 48 weeks were assessed with Wilcoxon signed-rank tests. Descriptive summaries are provided for efficacy and safety endpoints. Continuous variables are summarized by number of participants, median (IQR) and/or range unless otherwise indicated. Categorical variables are presented as number and percent. Bone turnover marker analyses are reported as both absolute and percent change from baseline.

## RESULTS

### Participants

Twelve participants were initiated on burosumab and completed 48 weeks of treatment. The median age was 16 years (range 6-38), with 5 adult and 7 pediatric participants (Table 1 and Suppl Appendix). At baseline all participants demonstrated low phosphate, TmP/GFR and 1,25-dihydoxyvitamin D levels, with elevated FGF23 levels. Participants had an overall high degree of FD burden and physical disability. Skeletal Burden Score is a measure of the proportion of the skeleton involved with FD, ranging from 0 (no FD) to 75 (panostotic FD)[24]; in this study the median Skeletal Burden Score was 64.5 (range 28-75)(Fig 1E and F, Suppl Appendix). Eight of the 12 participants required use of assistive ambulation devices, including wheelchairs, walkers, and/or crutches. Eleven participants had previously been treated with conventional therapy prior to the washout period. No pediatric participants had active rickets. Eleven of the 12 participants had MAS-associated endocrinopathies, which were managed medically throughout the trial according to standard guidelines [25]. One adult participant was diagnosed with hypercortisolism related to bilateral adrenal adenomas prior to initiation of burosumab. She was treated with bilateral adrenalectomy 7 weeks after starting burosumab and recovered without sequelae. Four adult participants received concomitant anti-resorptive treatment for management of FD-related bone pain, including 3 treated with denosumab, and 1 treated with pamidronate (Suppl Appendix). All anti-resorptives were initiated >1 year prior to burosumab treatment and were continued at the same dose throughout the trial. The final dose of burosumab ranged from 0.5 mg/kg every 4 weeks to 2.3 mg/kg every 2 weeks. Four of the 5 adult participants were transitioned to every 2-week dosing.

**Table 1.** Baseline Characteristics.

| Characteristic (n=12) | Median (IQR) or n (%) |
| --- | --- |
| Age (years) | 16 years (20.5, range 6-38) |
| Female/Male | 8 (67%)/4 (33%) |
| Skeletal Burden Score<sup>1</sup> | 64.5 (29) |
| Uses assistive ambulation device | 8 (67%) |
| McCune-Albright endocrinopathies | 11 (92%) |
| - Precocious puberty | - 10 (83%) |
| - Hyperthyroidism | - 5 (42%) |
| - Growth hormone excess | - 6 (50%) |
| - Hypercortisolism (neonatal-onset) | - 1 (8%) |
| - Hypercortisolism (adult-onset)<sup>2</sup> | - 1 (8%) |
| Phosphorus Z-score | -2.9 (1.7) |
| FGF23 (pg/mL) | 131 (176) |
| TmP/GFR | 0.76 (0.27) |
| 1,25-vitamin D (ng/mL)<sup>3</sup> | 21 (33) |
| 25-vitamin D (ng/mL)<sup>4</sup> | 38 (20.5) |
| PTH (pg/mL)<sup>5</sup> | 36.0 (39.1) |

<sup>1</sup>Semi-quantitative measure of the proportion of the skeleton involved with FD, ranging from 0 (no FD) to 75 (panostotic FD). See Collins et al, J Bone Min Res 2005 for details. <sup>2</sup>Participant was diagnosed with hypercortisolism prior to burosumab initiation and underwent bilateral adrenalectomy at week 7. <sup>3</sup>Normal reference range is 20-79 ng/mL. <sup>4</sup>All participants maintained levels ≥ 20 ng/mL throughout the study.
<sup>5</sup>Normal reference range is 15-65 pg/mL.

### Biochemistries

Serum phosphate levels increased into the target range in 12/12 (100%) of participants, increasing from a median of -2.88 (1.65) at baseline to 0.22 (1.37) at week 48 (Fig 2A and B). This was accompanied by improvements in 1,25 vitamin D levels from 20.5 (32.75) to 44 (43.75) ng/mL, and TmP/GFR from 0.76 (0.27) to 1.29 (0.43) mmol/L (Fig 2C and D).

**Figure 2.**
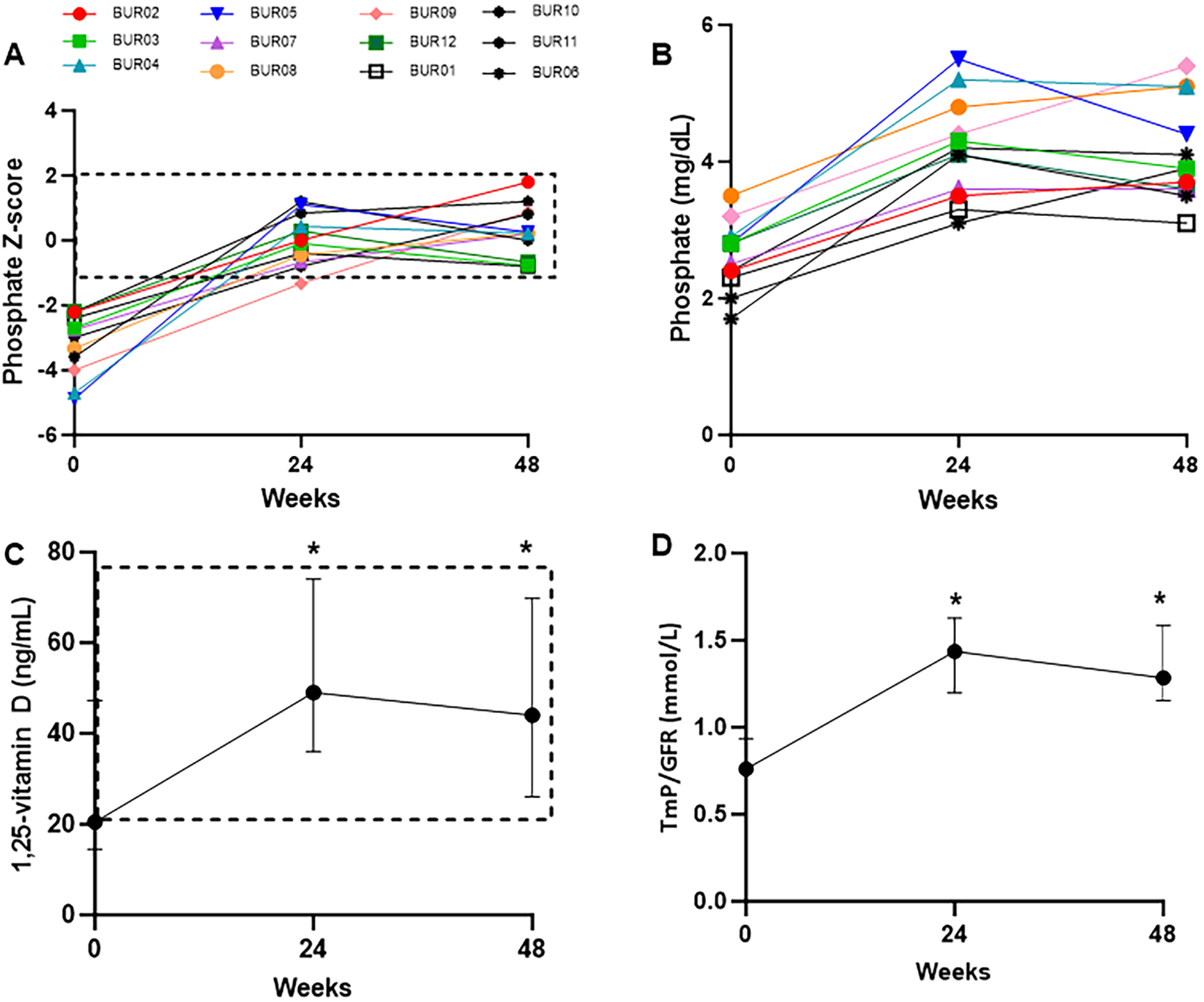
Biochemical changes in response to burosumab. A) 100% of participants achieved a serum phosphate level in the target range (age-and sex-adjusted Z-score between -1 and 2, dashed box) at week 48. B) Individualized changes in unadjusted serum phosphate levels. C) Changes in serum 1,25-vitamin D levels. Data are expressed as median and interquartile range. The dashed box indicates the normal range. *indicates significant change from baseline (p=0.006 for 24 weeks, 0.02 for 48 weeks, signed rank test). D) Changes in TmP/GFR. Data are expressed as median and interquartile range. Normal ranges vary by age and sex, with details available in Payne RB. Ann Clin Biochem. 1998. *indicates p=0.005 for both 24 and 48 weeks, signed rank test.

Changes in serum bone turnover markers are shown in Figure 3. Four participants had clinical variables which confounded bone turnover marker analyses, including concomitant denosumab treatment (participants BUR06, 10, and 11), and hypercortisolism treated with bilateral adrenalectomy at week 7 (BUR01). In the remaining participants, alkaline phosphatase levels were markedly elevated at baseline and showed consistent declines at weeks 24 and 48 of -41% (90) and -49% (96), respectively, representing a median decline of -364 (244.5) U/L from baseline. Osteocalcin levels showed an increase at week 24, with a reduction toward baseline levels at week 48. There were no consistent changes across the cohort in P1NP or CTX levels (Suppl Appendix).

**Figure 3.**
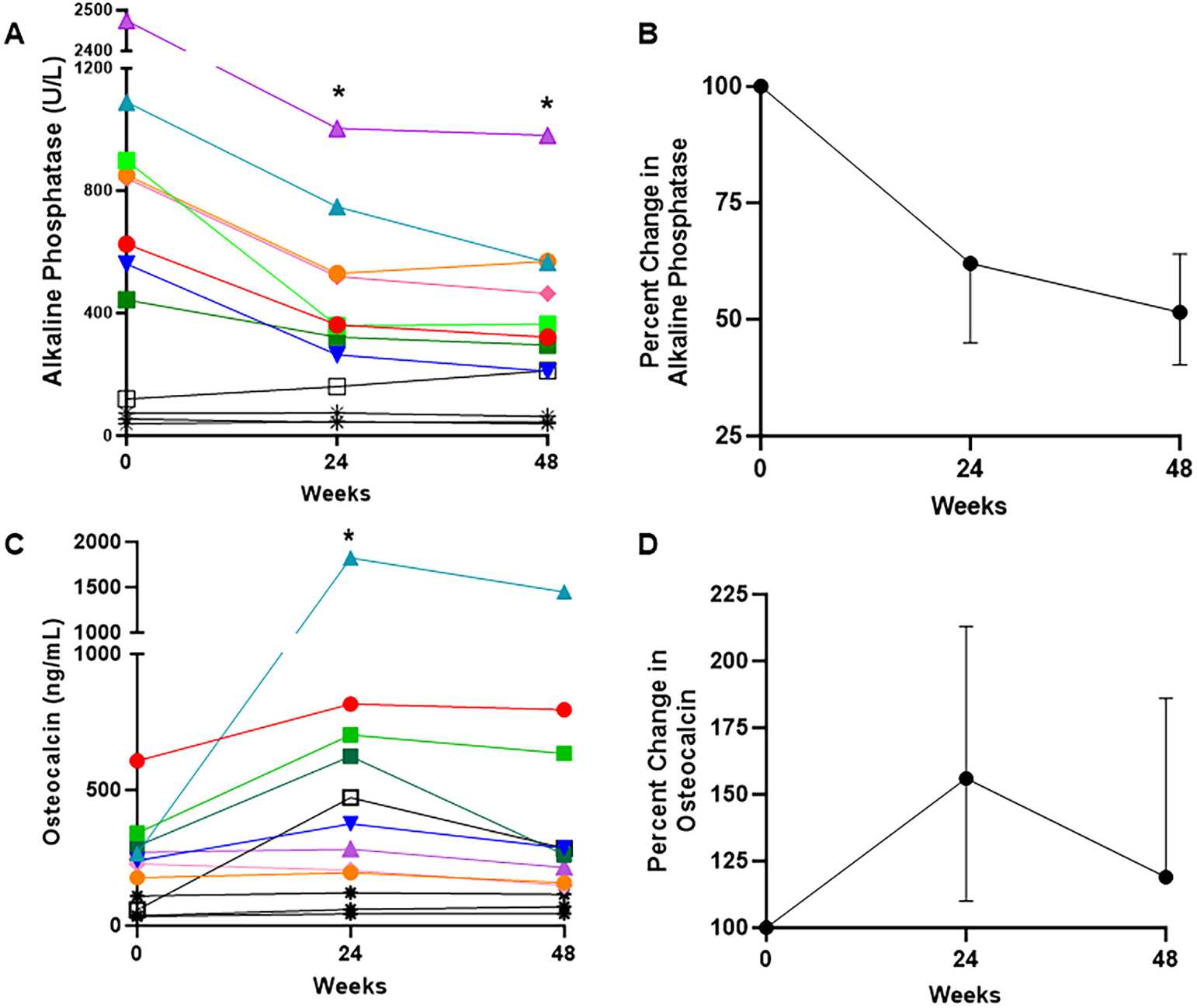
Bone turnover marker changes in response to burosumab. A) Individualized changes in alkaline phosphatase levels in the entire cohort (n=12). * indicates significant changes from baseline (p=0.008 for both weeks 24 and 48, signed rank test). Note that participants shown in black have confounding variables impacting bone turnover markers, including concomitant denosumab treatment (BUR10, 11, and 06, stars), and hypercortisolism status-post total adrenalectomy at week 7 (BUR01, open box). B) Percent change from baseline in alkaline phosphatase levels in the remaining cohort (n=8) are shown as median and interquartile range. C) Individualized changes in osteocalcin levels in the entire cohort (n=12). * indicates significant changes from baseline (p=0.04 for week 24, signed rank test). D) Percent change from baseline in osteocalcin levels in the remaining cohort (n=8) are shown as median and interquartile range.

### Functional and Patient-Reported Outcomes

PROMIS questionnaire scores in pediatric participants showed trends toward improvements in all domains at week 48, including reductions in Pain Intensity (p=0.06), Pain Interference (p=0.06), and Fatigue (p=0.15), and an increase in Mobility (p=0.12)(Fig 4A-D)(n=7). There were no consistent trends in PROMIS scores in adults (Fig 4E-H)(n=5). There was a potential trend toward improvement in SF10 physical health scores in pediatric participants (p=0.13), but no clear trends in SF36 physical domain scores in adults (Suppl Appendix).

**Figure 4.**
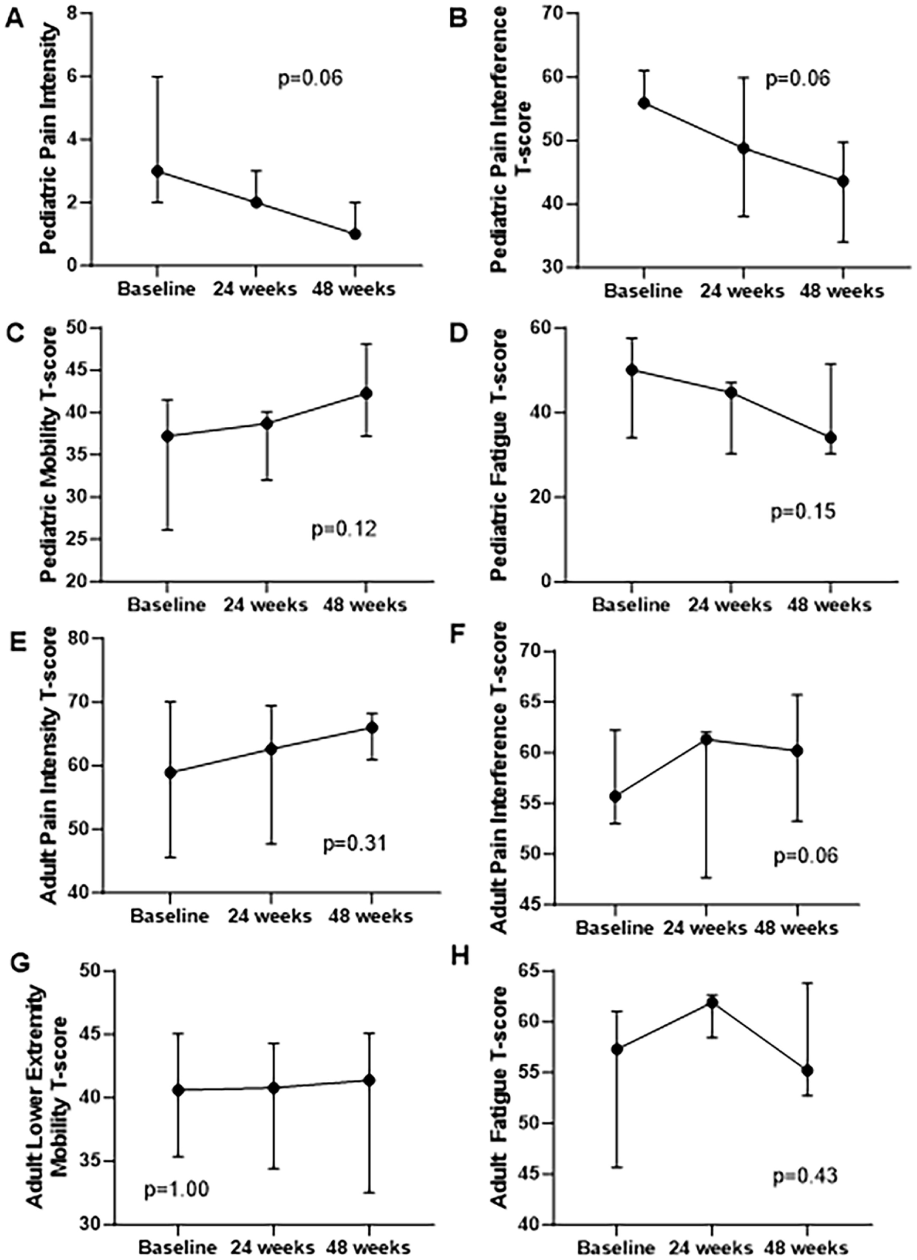
Change in PROMIS scores. Results for pediatric participants (n=7) are shown in the top four panels, which show consistent trends toward improvement in (A) pain intensity)(p=0.06, signed rank test), (B) pain interference (p=0.06, signed rank test), (C) mobility (p=0.12, signed rank test), and (D) fatigue (p=0.15, signed rank test). The lower four panels (E-H) show corresponding results for adult participants (n=5), which show no clear trends.

Two pediatric participants demonstrated transformational improvements in mobility after initiation of burosumab. BUR03 was a 10-14 year-old girl who had received conventional treatment for the previous 3 years. She became a full-time wheelchair user 3-years prior, and at her baseline visit was able to ambulate only 5-10 feet without assistance. By week 24 she was ambulating primarily without assistance. By week 48 she had achieved full-time independent ambulation and no longer required assistive devices for any activities. BUR06 was a 5-9 year-old girl who had received conventional treatment for the previous 4 years. She had never walked independently, and at her baseline visit was unable to perform any weight-bearing activities involving her lower extremities. By week 24 she was able to bear crawl (i.e. ambulate in a quadruped position) for mobility around her home. By week 48 she had progressed to using a walker at home and school, able to ambulate up to 50 feet.

For the qualitative Activities of Daily Living Assessment, participants and/or caregivers provided, in their own words, 3 activities of daily living that are impacted by FD at home or school/work (Fig 5A). There was a progressive increase in the proportion of participants reporting “much” or “very much” improvement in these activities between weeks 24 and 48 (Fig 5B).

**Figure 5.**
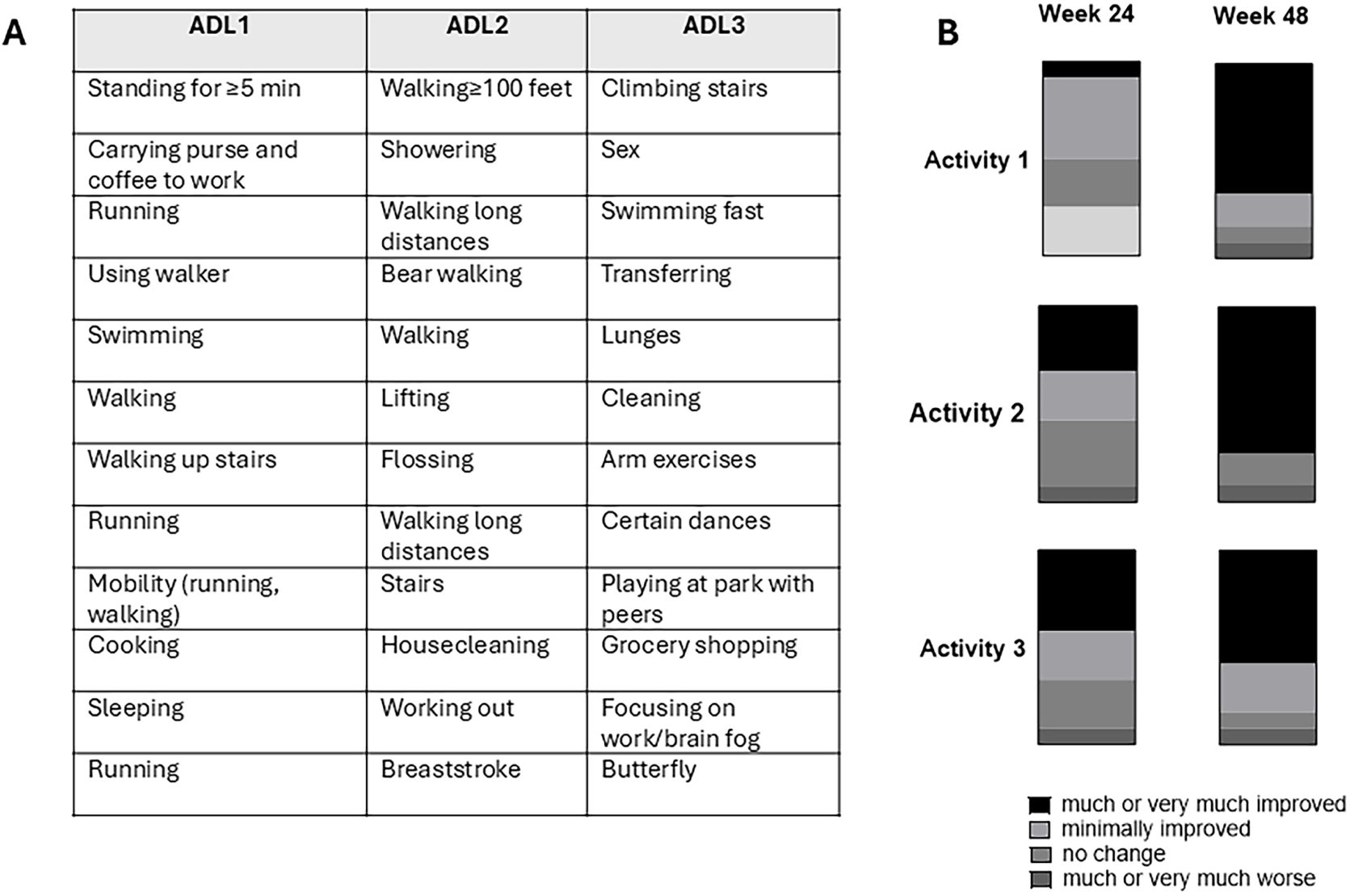
Change in qualitative Activities of Daily Living questionnaire. A) Each participant provided a list of 3 activities that are affected by their fibrous dysplasia. B) The proportion of participants reporting “much improvement” or “very much improvement” is shown in black, increasing between weeks 24 and 48.

No changes were noted in manual muscle testing or range-of-motion assessments between baseline and 48 weeks. Due to high levels of physical disability, only 4 participants were able to complete a 9-minute walk test, showing no changes.

### Safety Endpoints

Safety is summarized in Table 2. Nine out of 12 participants had at least one treatment-related adverse event. This included 8 incidences of hyperphosphatemia in 6 participants, classified as mild (grade 1) which resolved with resolved with pre-specified burosumab dose reductions according to the protocol. Three participants experienced a total of 25 injection site reactions, classified as mild (grade 1) and self-resolving. No adverse events resulted in treatment discontinuation.

**Table 2.** Safety Summary.

| Category | Number of events | Number of participants<br>n, % |
| --- | --- | --- |
| Total Adverse Events | 50 | 10 (83%) |
| Treatment-Related Adverse Events | 33 | 9 (75%) |
| - Hyperphosphatemia | - 8 | - 6<br>(50%) |
| - Injection site reactions | - 25 | - 3<br>(25%) |
| Serious Adverse Events | 2 | 2 (17%) |
| Serious Adverse Events Related to Treatment | 0 | 0 |
| Grade 3 or 4 Adverse Events | 1 | 1 (8%) |

There were no notable changes in safety labs, including calcium, PTH, urinary calcium, or eGFR (Suppl Appendix). Two participants had nephrocalcinosis at baseline, which remained stable at 48 weeks, and no additional participants developed nephrocalcinosis over the course of the study. No participants tested positive for anti-drug antibodies.

There were no notable changes in ^18^F-NaF PET/CT scans between baseline and 48 weeks (Suppl Appendix). FD lesion biopsies at baseline and 48 weeks were available from 4 adult participants, with no qualitative changes noted on histological evaluation. There were no quantitative differences in the cellularity or proportion of fibrous tissue (Suppl Appendix), suggesting that burosumab did not have a pro-proliferative effect on lesional tissue.

## DISCUSSION

Burosumab treatment in patients with FD was well-tolerated and associated with robust improvements in serum phosphate, pharmacodynamic measures, and trends toward patient-reported outcomes. Treatment was associated with restoration of ambulation in 2 severely affected children. These findings support targeting a high-normal phosphate level as a safe and effective treatment strategy in patients with FD.

Results from this trial address a critical knowledge and treatment gap for patients with FD relative to other FGF23 excess disorders. Previous studies have demonstrated safe and efficacious use of burosumab in the more common genetic disorder XLH [10 11 13 15], and the paraneoplastic disorder TIO [12 14]. However, unique considerations underscore the critical need to extend these investigations to the FD population. Because FD bone is inherently dysplastic and poorly mineralized, patients are prone to fractures and bowing deformities starting in early childhood [4-7], which are compounded by hypophosphatemia due to the vulnerability of dysplastic bone [8 24]. As a result, patients with FD and hypophosphatemia classically develop substantial skeletal-related morbidity, including early ambulation loss and progressive scoliosis [6 26](Fig 1). This results in an overall greater severity of disability compared to typical patients with XLH and TIO, where (apart from sequelae of hypophosphatemia) bone appears to be normally formed. [9 27 28].

Burosumab treatment in this study resulted in robust and expected improvements in phosphate and other pharmacodynamic measures, including 1,25-vitamin D and TmP/GFR. Alkaline phosphatase levels declined substantially by -49% (-364 U/L), exceeding results from trials in XLH and TIO, which ranged between -20 and -33% [14 15 29]. Bone turnover is a complex surrogate marker in FD, because altered differentiation of Gα_s_-mutation bearing osteoprogenitor cells results in high turnover even in the absence of hypophosphatemia [2 30]. Alkaline phosphatase levels correlate with overall FD burden, and have been historically used as a biochemical marker of lesional activity [22 24 30 31]. Levels typically remain elevated even after initiation of conventional therapy; for instance, participants did not exhibit substantial changes following the 2-week washout period, supporting a minimal effect of conventional therapy on this metric (Suppl Appendix). Findings from this study suggest that hypophosphatemia may contribute to a larger proportion of alkaline phosphatase elevation in patients with FD than previously recognized, pointing to the likely underappreciated contribution of hypophosphatemia to disease burden. These findings also highlight the need to consider how underlying pathogenic mechanisms impact biomarkers, particularly in mosaic diseases. The relative stability of P1NP and CTX levels over the course of this trial suggests they may better reflect high turnover due to FD lesional activity, while alkaline phosphatase may better reflect lesional mineralization.

Beneficial effects of burosumab were further supported by clinical observations. This study was not powered to detect statistical changes in patient-reported outcome tools; however, pediatric participants demonstrated consistent trends toward improvement in all domains. This is further supported by transformational mobility gains in 2 children. Because FD lesions progress during childhood, ambulation loss (if present) typically occurs during the pediatric period [32]. Substantial mobility gains after ambulation loss are therefore outside the expected disease course [26 32]. Several case reports of burosumab treatment in children with FD describe similar functional gains [33 34], although others are limited to biochemical improvements [35-37]. In this study, adult participants showed disparate results, with no trends toward improvement in PROMIS questionnaires or ambulation. Taken together, these findings support that early burosumab treatment may potentially mitigate the impact of hypophosphatemia on FD-related morbidity, including ambulation loss. However, compounding effects of long-term disability (such as joint contractures and deconditioning) are likely not reversible with improved mineral metabolism.

This study included a qualitative Activities of Daily Living questionnaire, with the goal of assessing individualized changes after burosumab treatment. Physical function tools used in other burosumab trials (such as Sit to Stand, walk tests, and jump tests) are not feasible in patients with severe physical disabilities; changes in self-reported ADLs were therefore investigated as a potential functional marker. While numerous standardized scales have been developed, they are primarily geared toward individuals with neurocognitive disorders [38 39], and do not encompass the broad spectrum of physical impairments that impact patients with FD across the lifespan. The instrument developed for this study is not a validated tool and results should be interpreted in that context; however, the self-reported activities provide insight into daily impacts of FD in individuals with high disease burden. The progressive self-reported improvements between weeks 24 and 48 are in concert with effects of burosumab on physical function in other populations [40-42], and provide further support for beneficial effects in patients with FD.

Results from this trial demonstrated a favorable safety profile. Adverse effects were mild and fell within expected parameters for burosumab use. While hyperphosphatemia occurred at a higher frequency compared to previous trials, it remained mild and was easily corrected with dose adjustments. Importantly, radiographic and histologic evaluations revealed no adverse effects on lesional activity, suggesting that blockade of FGF23 production from Gα_s_-mutation bearing osteoprogenitor cells is unlikely to increase FD malignancy risk or lesion progression.

This is the first study to investigate high-normophosphatemia as a therapeutic target for burosumab treatment. The trial design was informed by natural history data showing a linear relationship between serum phosphate and FD-related fractures and deformities, with higher complication rates in patients with both frank (age and sex-adjusted Z-score ≤-2) and low normophosphatemia (Z-score > -2 to ≤ -1) compared to those with high-normophosphatemia (Z-score -1 to 2) [8]. While few studies have investigated optimal phosphate targets in other FGF23 disorders, previous trials demonstrated clinical improvements upon achieving low-normophosphatemia in patients with both XLH and TIO [10-12 14]. International consensus guidelines in XLH and TIO therefore recommend titrating burosumab to achieve low-normal phosphate levels [16-18], although this remains an area of debate [43]. However, the increased vulnerability to hypophosphatemia and higher skeletal-related morbidity in patients FD with compared to other FGF23 excess disorders supports a more robust therapeutic target. Findings from this study demonstrate that high-normophosphatemia is a safe and effective treatment goal in FD, and further research is needed to define optimal targets in other FGF23 disorders. In particular, the mosaic RASopathy Cutaneous Skeletal Hypophosphatemia Syndrome shares phenotypic overlap with FD [44], and growing evidence suggests a similar approach may be beneficial [45-51].

Several limitations impact interpretation of this study. FD is a rare disorder, and patients with high disease burden and frank hypophosphatemia represent a small subset of the overall population. This limited the cohort size and precluded inclusion of a control arm. Because of their shared underlying pathophysiology, the trial was designed to include both children and adults. While this increases the generalizability of the results, it impacted interpretation of patient-reported outcomes (which include age-specific questionnaires) across the cohort. Three adults received concomitant denosumab treatment, which limited our ability to interpret mineral markers. Denosumab has become a standard treatment in adults with FD [23 52], particularly those with high disease burden [53]. We therefore elected to include participants receiving denosumab to ensure the study cohort reflected the overall target population of FD patients impacted by hypophosphatemia. It also is conceivable that co-treatment with burosumab may potentially enhance denosumab-induced lesional mineralization by ensuring adequate phosphate stores [54]. This trial provides the first safety and efficacy data in individuals co-treated with both therapies, and will inform the development of future trials and evidence-based treatment guidelines. Interpretation of biopsy specimens was another limitation. Dysplastic fibro-osseous tissue in FD does not consistently incorporate tetracycline labeling and cannot be analyzed with standard histomorphometric indices. Biopsies are also confounded by the mosaic nature of FD, leading to broad heterogeneity in lesion locations and composition. Biopsy specimens were therefore not feasible to investigate therapeutic effects of burosumab on lesional mineralization and were limited to pre-specified safety analyses.

Results from this trial demonstrate that burosumab treatment targeting high-normophosphatemia in patients with FD is safe, restores phosphate homeostasis, and improves bone turnover. Burosumab has the potential to lead to functional improvements and ambulation gains in severely affected children, and represents a valuable tool to reduce the impact of FD-related disability. These findings support burosumab as a preferred approach to achieve normophosphatemia in patients with FD.

## Data Availability

All data produced in the present study are available upon reasonable request to the authors

