## Supplementary material for "A phase 2 trial of burosumab for treatment of fibroblast growth factor-23 mediated hypophosphatemia in children and adults with fibrous dysplasia": Suppl Appendix

### Figure S1. Overall trial design

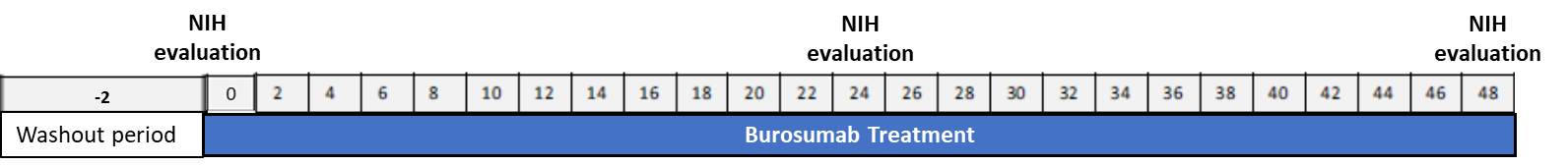

Participants were treated with burosumab for 48 weeks following a 2-week washout period. Evaluations were performed at the NIH Clinical Center at baseline, 24, and 48 weeks. Between visits burosumab was administered locally, and participants were monitored remotely with outpatient labs.

### Figure S2. Clinical flowchart

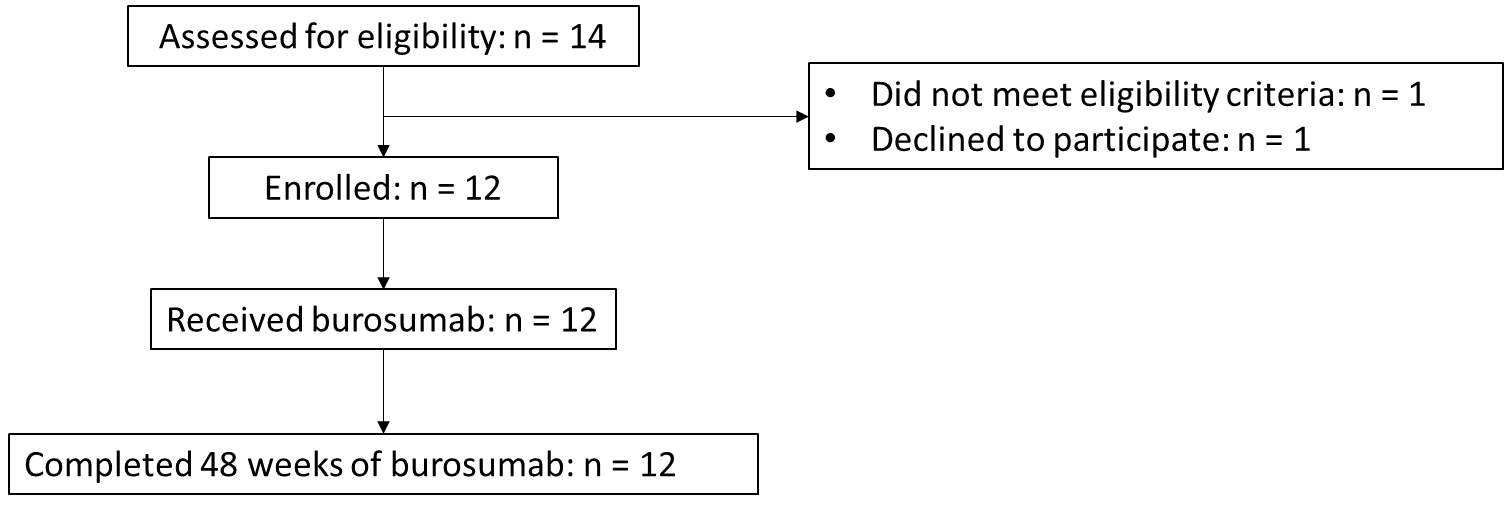

### Table S1. Individual participant characteristics

| **ID** | **Age (years) & Sex** | **Skeletal Burden Score^1^** | **MAS-associated endocrinopathies** | **Concomitant anti-resorptive treatment** | **Previous conventional therapy** | **Burosumab regimen at study completion** |
| --- | --- | --- | --- | --- | --- | --- |
| BUR01 | 30-39,F | 68 | Hypercortisolism (adult-onset, resolving), precocious puberty, hyperthyroidism, growth hormone excess | None | Yes | 0.7 mg/kg every 2 weeks |
| BUR02 | 20-29,F | 75 | Precocious puberty | Pamidronate^4^ | Yes | 0.7 mg/kg every 2 weeks |
| BUR03 | 10-14,F | 61 | Precocious puberty, hyperthyroidism, growth hormone excess | None | Yes | 0.8 mg/kg every 2 weeks |
| BUR04 | 5-9,F | 75 | Precocious puberty, hyperthyroidism, growth hormone excess, hypercortisolism (neonatal-onset, resolved) | None | Yes | 2.3 mg/kg every 2 weeks |
| BUR05 | 10-14,F | 35 | Precocious puberty | None | Yes | 0.5 mg/kg every 2 weeks |
| BUR06 | 20-29,F | 43 | Precocious puberty | Denosumab^5^ | Yes | 1.0 mg/kg every 2 weeks |
| BUR07 | 15-19,M | 75 | Precocious puberty, hyperthyroidism, growth hormone excess | None | Yes | 0.8 mg/kg every 2 weeks |
| BUR08 | 10-14,M | 51 | Hyperthyroidism, growth hormone excess | None | Yes | 0.4 mg/kg every 2 weeks |
| BUR09 | 5-9,M | 69 | None | None | Yes | 1.5 mg/kg every 2 weeks |
| BUR10 | 30-39,F | 69 | Precocious puberty | Denosumab^6^ | Yes | 0.5 mg/kg every 2 weeks |
| BUR11 | 30-39,F | 28 | Precocious puberty | Denosumab^6^ | Yes | 0.5 mg/kg every 4 weeks |
| BUR12 | 15-19,M | 48 | Precocious puberty, growth hormone excess | None | No | 0.3 mg/kg every 2 weeks |

^1^Semi-quantitative measure of the proportion of the skeleton involved with FD, ranging from 0 (no FD) to 75 (panostotic FD). See Collins et al, J Bone Min Res 2005 for details.

### Figure S3. Bone turnover markers

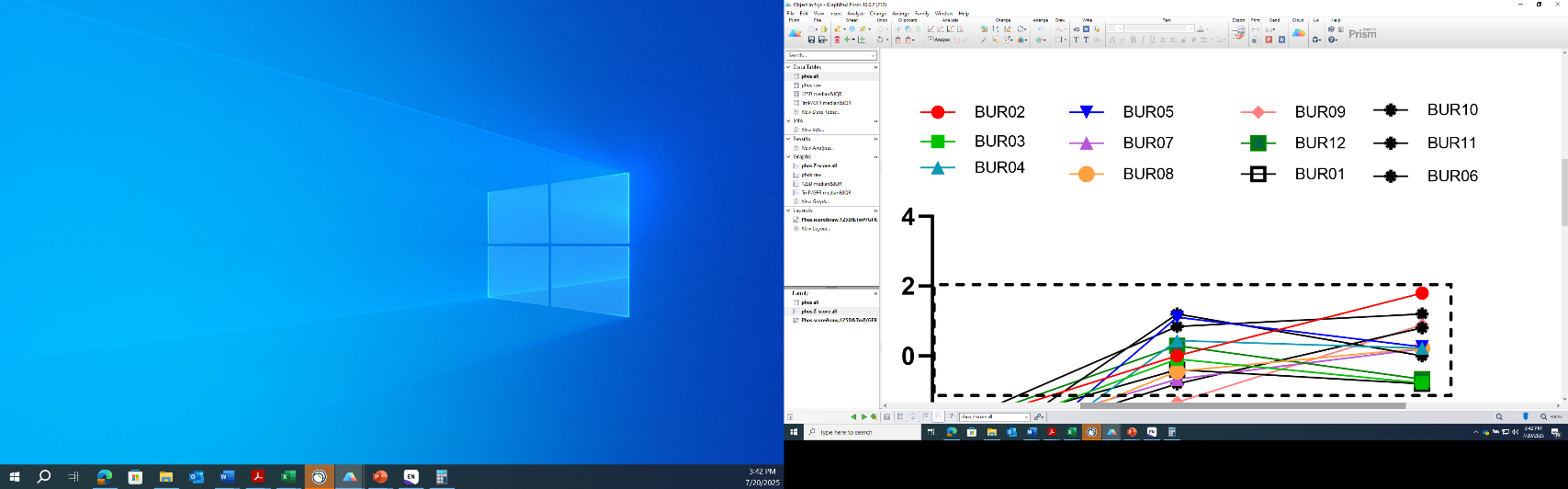

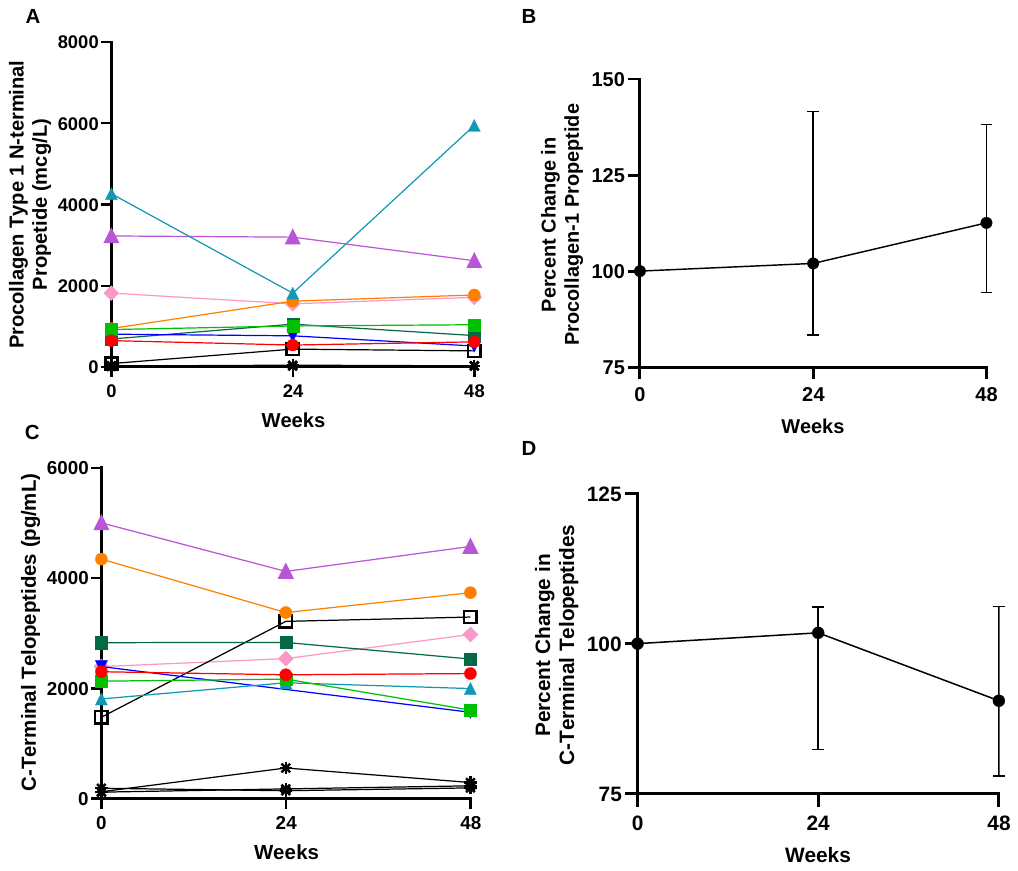

Individualized changes in procollagen-1 propeptide (A) and C-terminal telopeptides (C) in the entire cohort (n=12). Note that participants shown in black have confounding variables impacting bone turnover markers, including concomitant denosumab treatment (BUR10, 11, and 06, stars), and hypercortisolism status-post total adrenalectomy at week 7 (BUR01, open box). Percent change from baseline in procollagen-1 propeptide (B) and C-terminal telopeptides (D) in the remaining cohort (n=8) are shown as median and interquartile range.

### Figure S4. Additional Patient-Reported Outcomes

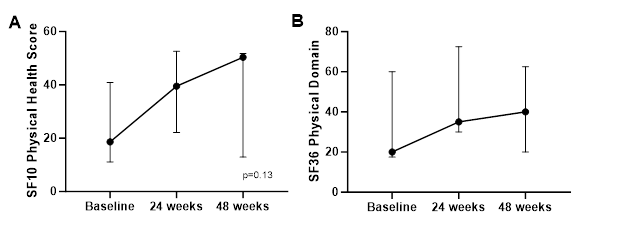

Data are expressed as median and interquartile range. Panel A shows a potential trend toward improvement in SF10 physical health scores in pediatric participants (p=0.13, signed rank test, n=7). Panel B showed results for SF36 physical domain scores in adult participants, with no clear trend (n=5).

### Table S5. Activities of Daily Living Questionnaire

| Complete items 1-3 at Baseline Visit | | Indicate level of change since Baseline on extent of impact | |
| --- | --- | --- | --- |
|  |  | Complete items 4-6 at Week 24 | Complete items 7-9 at Week 48 |
| Please list up to 3 activities of daily living that are affected by FD/skeletal features at home or school/work (e.g., mobility, climbing stairs, dressing, playing with peers, catching a bus); and estimate the extent of impact | 1) Activity:  Stops activity completely  Extremely limited  Moderately limited  Slightly limited | 4) Since last visit  Very Much Improved  Much Improved  Minimally Improved  No Change  Minimally Worse  Much Worse  Very Much Worse | 7) Since baseline  Very Much Improved  Much Improved  Minimally Improved  No Change  Minimally Worse  Much Worse  Very Much Worse |
|  | 2) Activity:  Stops activity completely  Extremely limited  Moderately limited  Slightly limited | 5) Since last visit  Very Much Improved  Much Improved  Minimally Improved  No Change  Minimally Worse  Much Worse  Very Much Worse | 8) Since baseline  Very Much Improved  Much Improved  Minimally Improved  No Change  Minimally Worse  Much Worse  Very Much Worse |
|  |  | N/A No activity listed in 2 | N/A No activity listed in 2 |
|  | 3) Activity: | 6) Since last visit | 9) Since baseline |
|  | Stops activity completely  Extremely limited  Moderately limited  Slightly limited | Very Much Improved  Much Improved  Minimally Improved  No Change  Minimally Worse  Much Worse  Very Much Worse | Very Much Improved  Much Improved  Minimally Improved  No Change  Minimally Worse  Much Worse  Very Much Worse |
|  |  | N/A: No activity listed in 3 | N/A: No activity listed in 3 |

### Figure S5. Safety labs

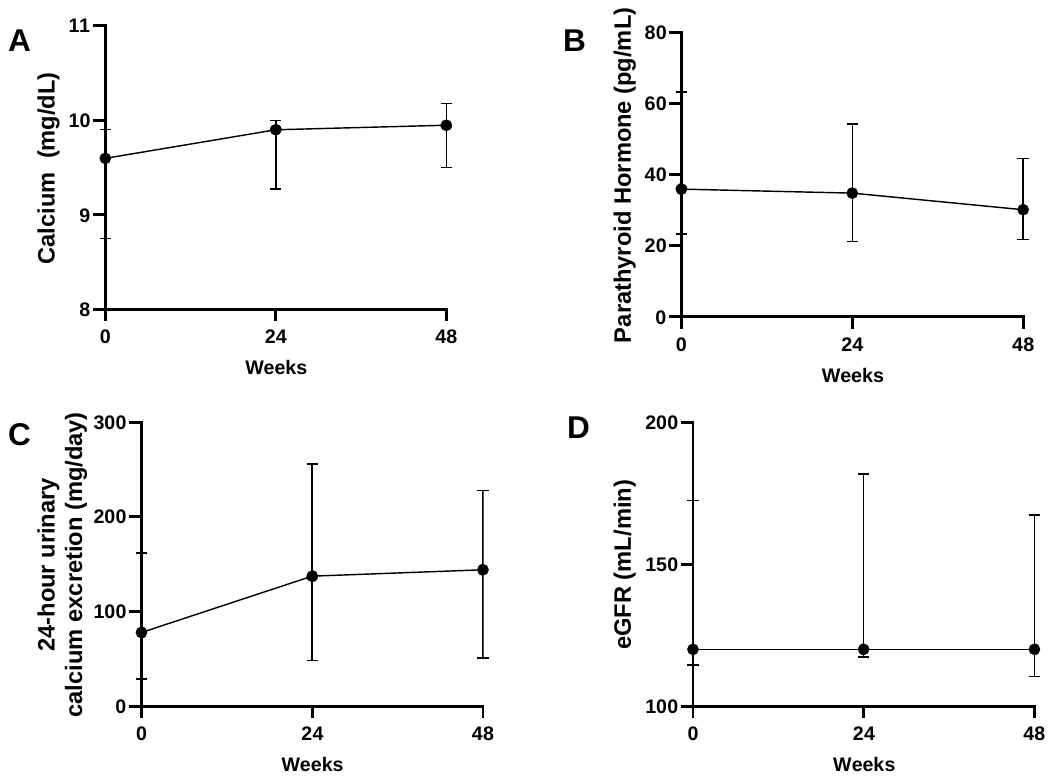

Data are presented as median and interquartile range.

### Figure S6. Markers of lesion activity

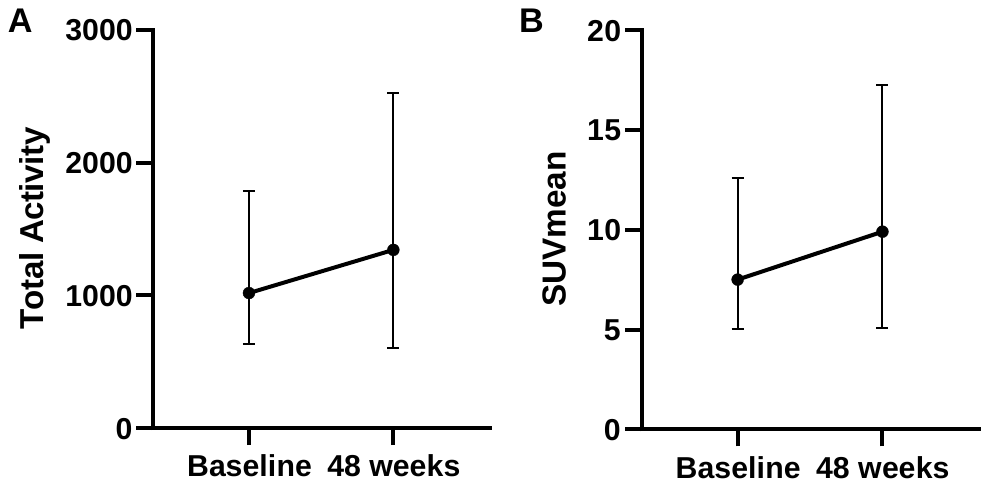

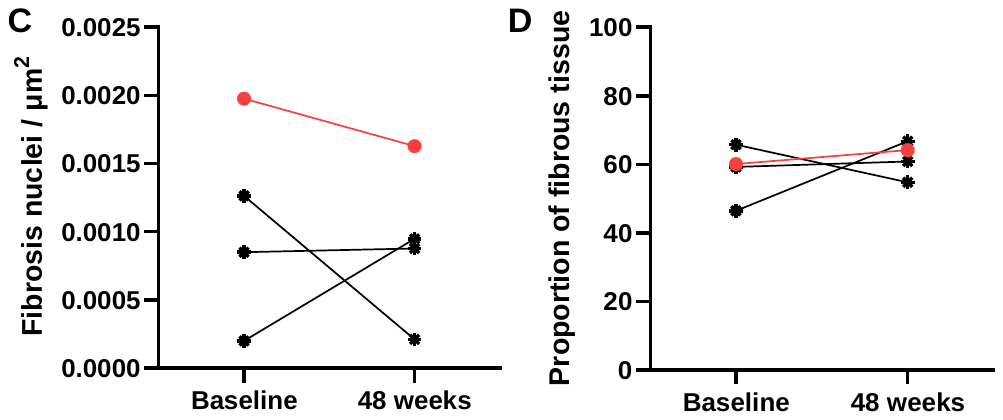

^18^F-NaF PET/CT metrics total activity (A) and SUV mean (B) are shown in median and interquartile range for the entire cohort (n=12). Histologic evaluation of pre- and post-treatment biopsies 4 adult participants (BUR02, 06, 10, and 11) show no quantitative differences in the cellularity (C) or proportion of fibrous tissue (D).

### Figure S7. H&E stains of biopsies before and after burosumab treatment

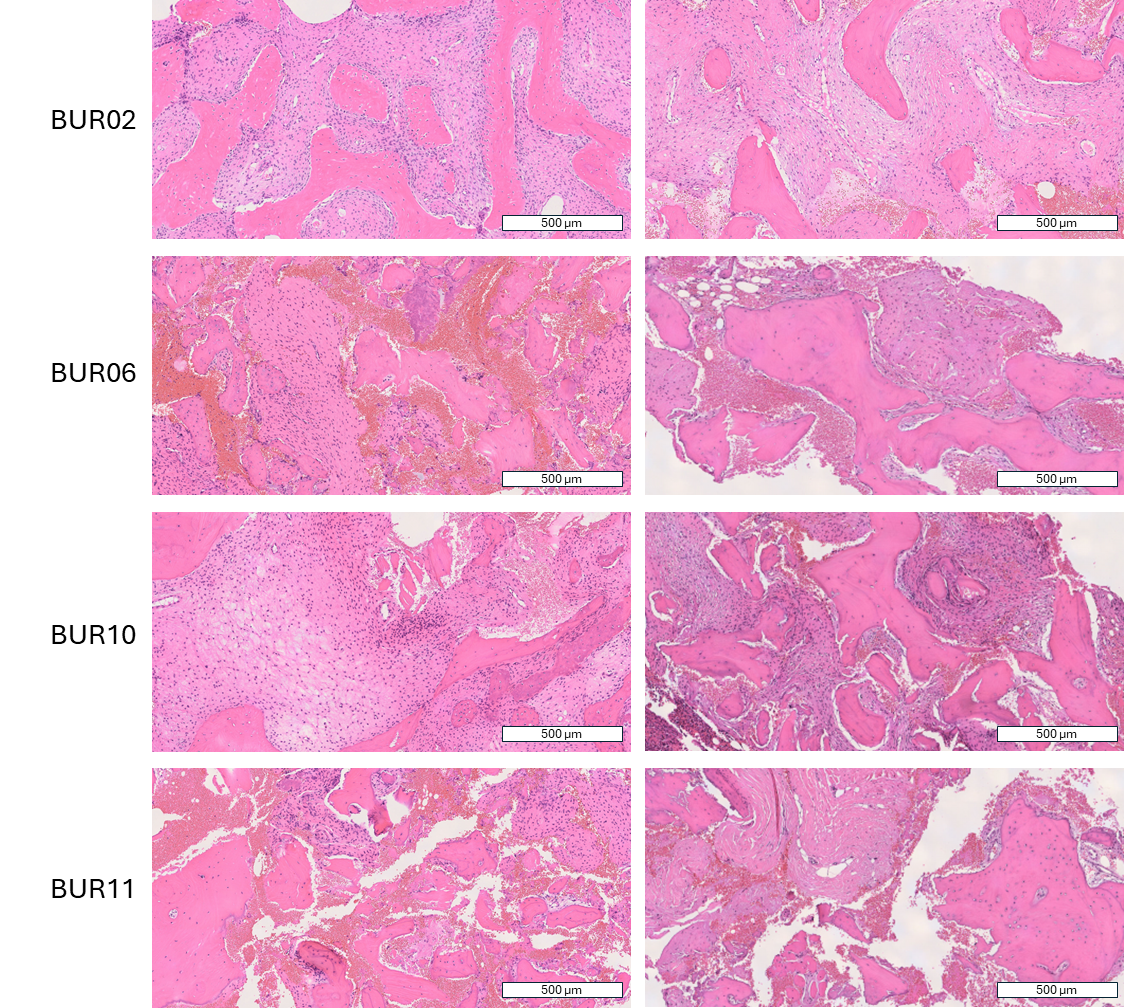

Low magnification images of a cross-section of all biopsies obtained from participants at baseline and 48 weeks. Locations included iliac crest (BUR02, 06, 10) and rib (BUR11).

### Figure S8. Alkaline phosphatase levels in participants previously treated with conventional therapy

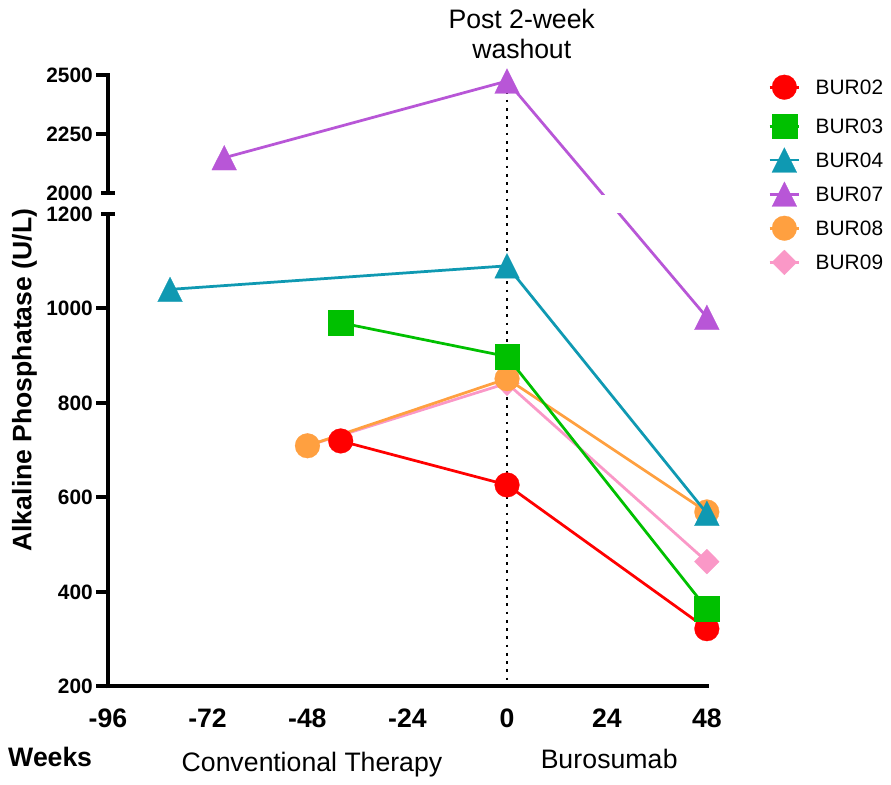

Retrospective review of alkaline phosphatase levels available for the subset of participants who were treated with conventional therapy prior to burosumab.
